# Primary care access and feasibility of Spatial Accessibility Index (SAI) in assessing primary care in rural Sri Lanka

**DOI:** 10.1101/2024.01.10.24301140

**Authors:** Parami Abeyrathna, Suneth Buddhika Agampodi, Manjula Weerasinghe, Shyamalee Samaranayake, Pahala Hangidi Gedara Janaka Pushpakumara

**Affiliations:** Department of Family Medicine, Faculty of Medicine and Allied Sciences, Rajarata University of Sri Lanka, Anuradhapura, Sri Lanka; Department of Internal Medicine, School of Medicine, Yale University, Connecticut, United States of America; Department of Community Medicine, Faculty of Medicine and Allied Sciences, Rajarata University of Sri Lanka, Anuradhapura, Sri Lanka; Department of Family Medicine, Faculty of Medical Sciences, University of Sri Jayawardhanapura, Nugegoda, Sri Lanka

**Keywords:** primary care, spatial accessibility of health services, healthcare disparities, healthcare planning

## Abstract

Despite decades of global health efforts and policies, inequitable access to primary care in low and middle-income countries remains an ongoing challenge. This study assessed accessibility to curative sector-primary healthcare services in the Anuradhapura district of Sri Lanka in 2020 and 2021. In this study, we included both private and state-owned primary care facilities (PCF) and their corresponding primary care doctors (PCD), covering both allopathic and complementary and alternative medicine (CAM) practices. Spatial locations of private sector PCF of the entire district were collected by conducting a comprehensive road-based survey and documenting information from publicly displayed boards. Data on state PCF were collected from provincial health departments (allopathic and Ayurveda). The Spatial Accessibility Index (SAI) of 657 GND (Grama Niladhari Division: the smallest administrative division) was calculated in ARC-GIS using the 2-Steps-Floating-Catchment-Area method in 5 km and 10 km spatial-distances from a GND. The majority of the PCF were allopathic (n=318, 78%) and belonged to the private sector (n=317, 79%) whereas CAM PCD represented one-fifth of the primary care workforce. Average SAI of 4.50X 10^-4^ and 4.67X 10^-4^ were reported for both 5 km and 10 km spatial-distance. A statistically significant difference in mean SAI was measured between urban and rural populations. The highest SAI were observed from urban populations and it gradually declined towards the rural remotes. The national primary care coverage target of 1 allopathic PCD per 5000 population was achieved only in 64% of the population in a 5 km spatial-distance and it further depleted to 25% with only of state allopathic PCD. SAI was negatively associated with the poverty headcount indices and distance to main roads and the main town (Anuradhapura). While the private allopathic sector is the main primary care provider in this study setting, a significant proportion of the population reported limited primary care access dueto their geographic and economic status. Health policymakers need to address primary care workforce shortages in rural for the planning of human and physical resources. SAI can be recommended as a feasible method for healthcare planning and assessing service coverage in low-facility settings.

## Introduction

Universal health coverage (UHC) refers to “all people having access to the full range of quality health services they need, when and where they need them, without financial hardships” (1). UHC depends on an equitable primary health care (PHC) system. In many countries, community and healthcare facility-based or primary care-oriented approaches integrated to deliver a more efficient and sustainable package of PHC. A primary care doctor (PCD) is the first contact personnel of a health system delivering continuous, comprehensive and coordinated care to patients for a minimum cost. Good primary care access has proven to reduce mortality by both communicable and non-communicable diseases (2,3) which were among the top 10 causes of death worldwide (4).

Disparities in primary care access affect UHC because of the unavailability of essential healthcare services and doctors and, increased out-of-pocket expenditures by indirect medical costs for traveling. According to the World Health Organization (WHO) and World Bank, half of the world’s population lacked access to essential healthcare services in year 2017 (5). Significant inequalities in the geographic distribution of primary care services were still observed from high-income countries like USA (6) and China (7). Locations of population in urban and rural territories, the extent of infrastructure development and poor socioeconomic status were among the main determinants for the observed disparities (8,9). Although WHO recommends a minimum of 80% population coverage of essential healthcare services through UHC (1), significant proportions of populations especially from lower-middle-income countries (LMIC) still report limited access (9,10). In South-East Asia, UHC service coverage indices (SCI) were ranged from 46% to 82% and Sri Lanka was placed in between: 66% (1).

Sri Lanka is considered a LMIC (10) with good maternal and child healthcare indicators over the past years (11). It has a pluralistic health system in which a mixture of medical practitioners from allopathic, Ayurveda, Yunani, Siddha, Homeopathic and indigenous medicine are involved in PHC delivery (11). The Ministry of Health (MoH) in Sri Lanka has proposed PHC reforms intending to achieve UHC by the year 2030 and highlighted the current issues in primary care services (12). Despite the national-level household surveys on healthcare utilization (13), primary care access has not evaluated in the Sri Lankan context previosly. The UHC-SCI is calculated weighing the international health regulation core scores, hospital bed densisty and healthcare workforce at country levels (9) and does not exclusively consider the spatial nature of healthcare access. The spatial accessibility is considered an accurate methord for evaluation of healthcare access nowadays with the use of geographic information system (GIS) technology (11). The current study was conducted to assess the spatial accessibility of curative sector primary care services in the Anuradhapura district of Sri Lanka.

## Methods

### Study setting

This study in the Anuradhapura district of Sri Lanka has conducted between August 2020 and December 2021. The district consisted of 22 divisional secretariat (DS) divisions and is primarily rural, with 96% of its population totaling 953,162 residents(12). These DS divisions has subdivided into 694 Grama Niladhari Divisions (GND), the smallest public administrative unit defined by their geographic areas. The majority of the population in Anuradhapura, as in the rest of Sri Lanka, relies on the allopathic healthcare system for their medical needs (13). Primary care services are provided by both the state and private sectors. Primary Medical Care Units (PMCU), Divisional Hospitals (DH), Outpatient Departments (OPD) of Base Hospitals (BH) and OPD of the Teaching Hospital (TH) serve as the main state sector primary care facilities (PCF) providing curative services in Anuradhapura (14). The private sector for curative care comprises private hospitals with OPD services and private sector allopathic practitioners, often referred to as general practitioners or GP, who are state medical officers practicing outside regular working hours (15). Preventive PHC services are provided through a state services consisting of a team led by the Medical Officer of Health (MOH). PHC services through CAM practices are provided through a network of state funded Ayurveda hospitals (16). Private PCF of CAM included several other practitioners following indigenous medicine, Yunani, Siddha, and homeopathy scattered in the remotes and sub-urban regions of Anuradhapura (17).

Despite the state PCF being listed, mapped, and included in public databases, the situation is different for private PCFs in Sri Lanka. While there is a registration system in place, managed by the regional director of health services, it is incomplete, and crucial data on the total number of facilities and their spatial locations are unavailable.

### Data sources and collection

All data collected for this analysis was either secondary data available in registers, publically displayed data or GIS data on PCF. Data on PCF and Primary Care Doctors (PCD) were collected from different sources (Table 1).

**Table 1.**
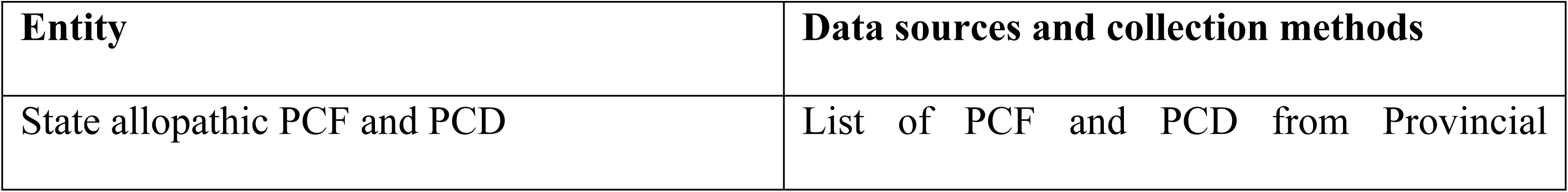
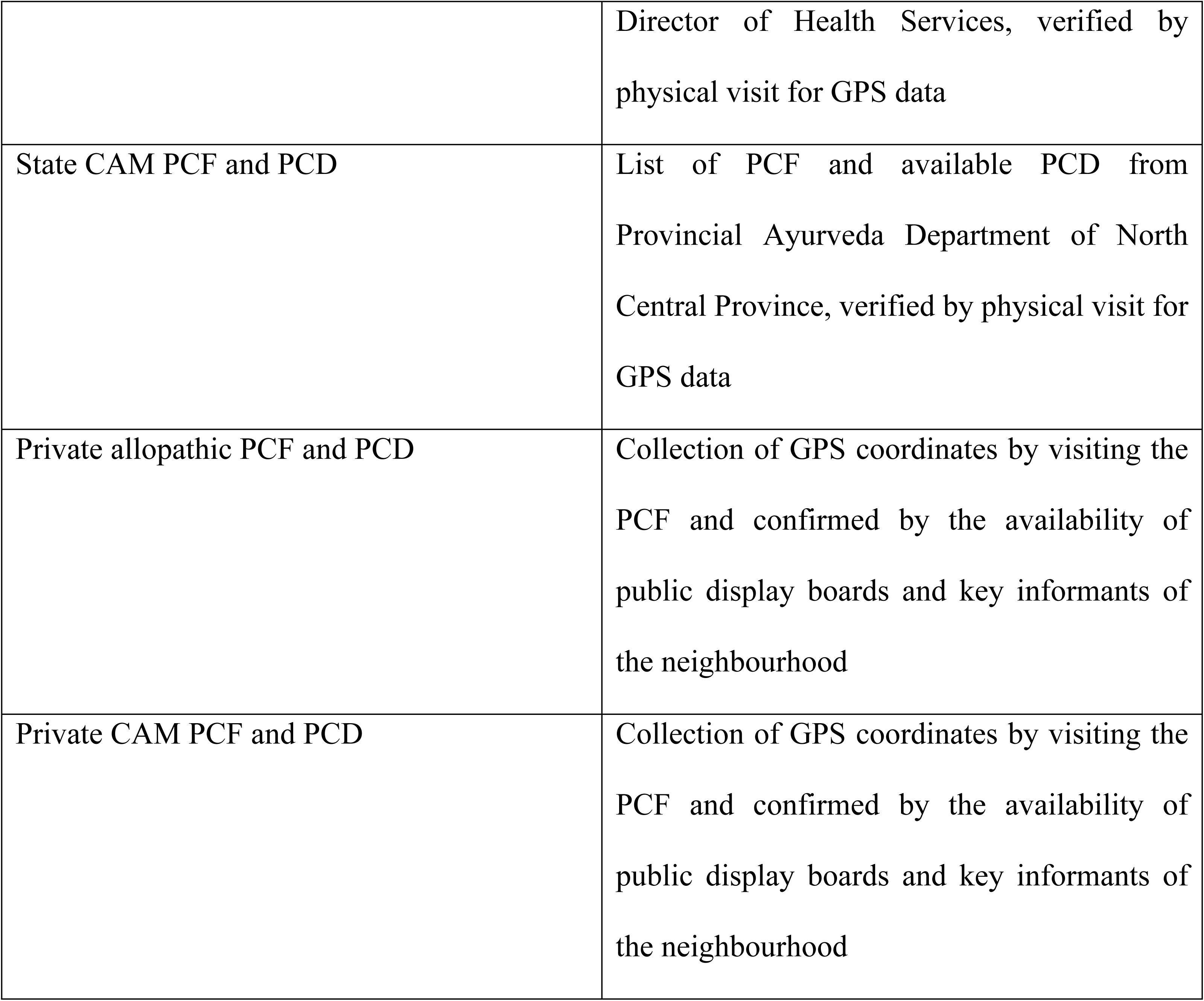
Data sources and collection methods for Primary Care Facilities (PCF) and Primary Care Doctors (PCD)

The total number of state allopathic and CAM PCD was referred from the official records from the Provincial Director of Health Services office and the Provincial Ayurvedic Department of the district. The global positioning system (GPS) locations of state allopathic and CAM PCF were verified with the site vists after referring to available records at these institutions.

The locations of private allopathic PCF were included only if they had a registration number from the Sri Lanka Medical Council (SLMC) on public display. Similar criteria were implemented for recognizing Private CAM practitioners who were registered at Ayurvedic Medical Council of Sri Lanka. The presence of a PCF was confirmed by the availability of public display boards and verification by a key informant who is well aware of the area such as *Grama Niladhari* or an officer of community-based organization. The locations of private allopathic and CAM PCF in the entire district were collected using the GPS coordinates by trained research assistants. GPS data were marked by using a special GPS device: Juno device (18). The spatial location, the no of Primary Care Doctors (PCD) at the facility and type of PCF were recorded and no information on individually identifiable data of the PCD was included.

Population data in the year 2019 of GND and divisional secretariat divisions were collected from official records (19). Spatial administrative maps of GND, divisional secretariat divisions and towns including road networks of the district were collected from the Department of Survey, Sri Lanka. Socio-demographic data were collected from the District Statistical Handbook of the Department of Census and Statistics of Sri Lanka (12).

### Data analysis

The Two Steps Floating Catchment Area-2SFCA (20) method was used to calculate the Spatial Accessibility Index (SAI) using ARC ESRI Geographic Information System (GIS) software version 9.5. A PCD to population ratio within areas of 5 km and 10 km spatial-distance from a PCF was calculated. SAI was calculated by summing up the PCD to population ratios that lie within the 5 km and 10 km boundary marked from the population center of each GND through ARC GIS software (21). SAI is indicated in terms of the number of PCD per 10,000 population within the particular spatial-distance.

The collected data of spatial maps with SAI were exported as Microsoft Excel sheets and analysis was conducted using IBM SPSS Statistics for Windows, Version 25.0. The nearest facility tool and nearby tables of ARC GIS were used to identify the nearest PCF, spatial distance to the nearest road with public transport, and spatial distance to Anuradhapura city from each GND. The Anuradhapura town, the only urban council of the district with 25 GND (22), was marked as the urban area of the district.

### Ethics statement

The ethics approval study was obtained from the Ethics Review Committee, Faculty of Medicine and Allied Sciences, Rajarata University of Sri Lanka (ERC/2020/66). The secondary data were obtained from the relevant authorities based on the administrative permission given by the authorised officers. No incentives were offered to any individuals participated in the study. All the participants who provided confirmation on locations of PCF were provided with a information leaflet and data were collected after obtaining the written informed consent. No personal identification or contact details were collected.

## Results

In the State Allopathic sector, primary care services was predominantly delivered by DH Type C, which housed 22 PCF (5.5% of the total) and 44 PCD (Table 2). Nevertheless, private allopathic providers played the central role, constituting 63.7% of PCF and 49% of PCD. The State Ayurvedic Sector contributed with 25 PCF (6.2%) and 56 PCD (11%), while the Private Sector Complementary and Alternative Medicine (CAM) included 60 PCF (14.8%) and 60 PCD (11%). In total, the district had 403 PCF and 523 PCD. These findings illuminate the diverse landscape of primary care provision, with a notable presence of private and CAM practitioners, offering valuable insights for healthcare planning and resource allocation in the region.

**Table 2.** An Overview of primary care facilities and primary care doctors in Anuradhapura District, Sri Lanka.

| Primary care facility | No of facilities (%) | No of primary care doctors (%) |
| --- | --- | --- |
| **State allopathic sector** |  |  |
| Teaching hospital of Anuradhapura | 1 (0.3%) | 20 (4%) |
| Base hospital | 6 (1.5%) | 30 (6%) |
| Divisional hospital-A | 2 (0.5%) | 8 (2%) |
| Divisional hospital-B | 9 (2.2%) | 27 (5%) |
| Divisional hospital C | 22 (5.5%) | 44 (8%) |
| PMCU | 21 (5.2%) | 21 (4%) |
| **Private allopathic** |  |  |
| Private allopathic | 257 (63.7%) | 257 (49%) |
| **State Ayurvedic sector** |  |  |
| State Ayurveda hospitals with OPD and MOH centers | 25 (6.2%) | 56 (11%) |
| **Private sector Complementary and Alternative Medicine (CAM)** |  |  |
| Private CAM practitioners | 60 (14.8%) | 60 (11%) |
| **Total** | **403(100%)** | **523(100%)** |

OPD: Out Patient Departments PMCU: Primary Medical Care Units MOH:Medical Officer of Health

Most of the PCF were close to urbanized regions and the suburbs in Anuradhapura district (Fig 1). PCF located close to the district border were scarce in number and scattered throughout.

**Fig 1.**
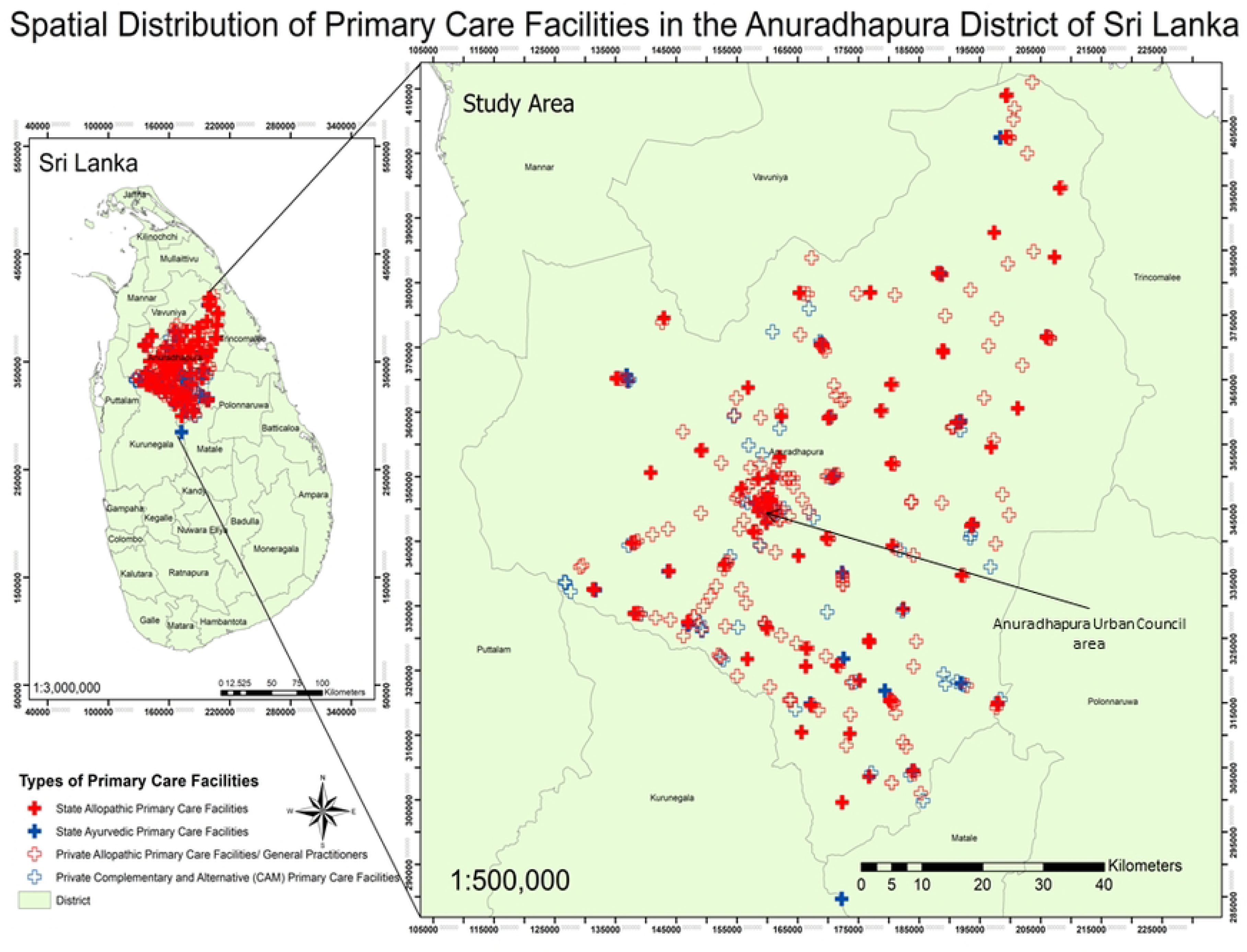
Spatial distribution of primary care facilities in Anuradhapura district of Sri Lanka. A total of 3 GND reported the highest SAI (15 /10,000) and located within a 5 km spatial-distance of Anuradhapura town (Table 2). A private allopathic PCF was closest to 377 GND (57%). Across all PCF, there is an average of 5 PCD per 10,000 population, with a mean spatial-distance of 2.4 km and a standard deviation of 1.8 km (Table 3). Urban areas exhibit higher PCD density (13 per 10,000 population) and shorter mean spatial distances (0.5 km at 5 km spatial-distance and 0.3 km at 10 km spatial-distance) compared to rural areas (4 doctors per 10,000 population and a mean distance of 2.5 km, SD = 1.8 km). The Allopathic state sector and the Allopathic private sector both report 2 PCDs per 10,000 population and similar mean distances. CAM PCF have 1 doctor per 10,000 population, with a mean spatial-distance of 2.3 km.

**Table 3.**
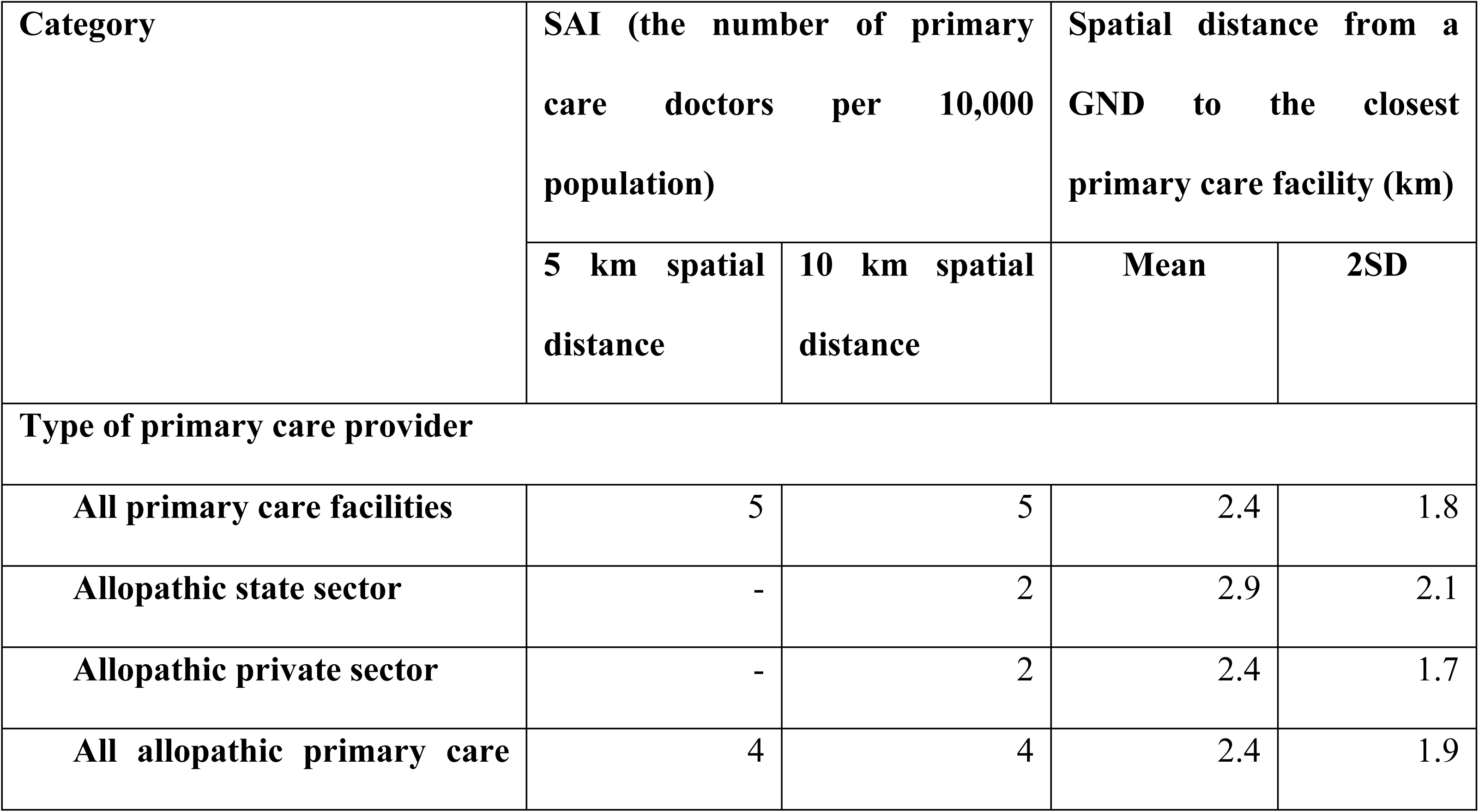
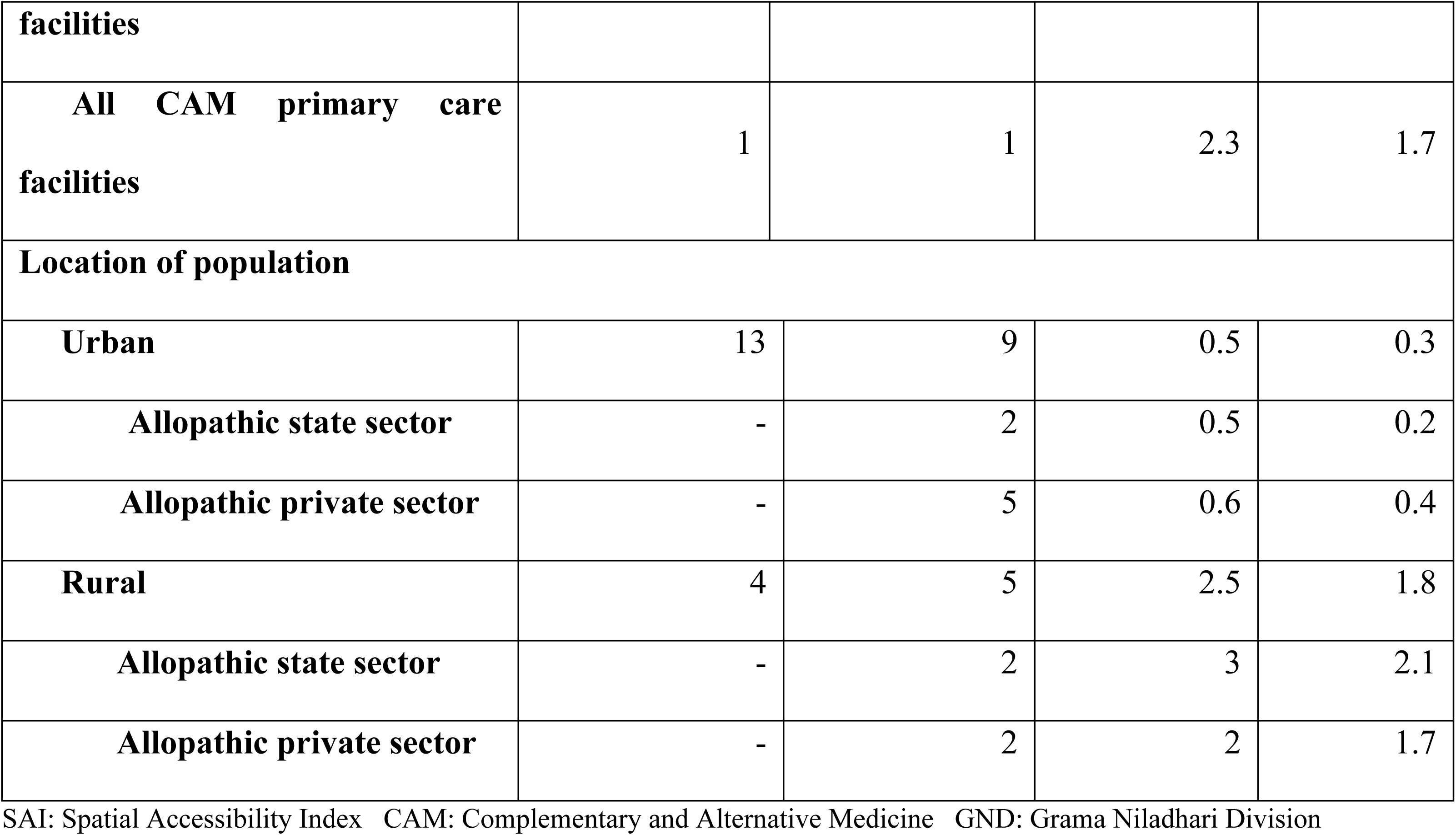
Spatial Accessibility Indices and proximity of curative sector primary care services in Anuradhapura district, Sri Lanka.

The spatial maps with SAI showed the inequitable distribution of SAI of primary care in the district between different types of providers and within different spatial distances from a GND (Fig 2). Higher SAI were observed in and around urbanized areas (Fig 1.). Lower values were more towards rural areas and district border. One-fourth of the population lived in areas with low primary care accessibility (SAI< 1.72 X 10^-4^).

**Fig 2.**
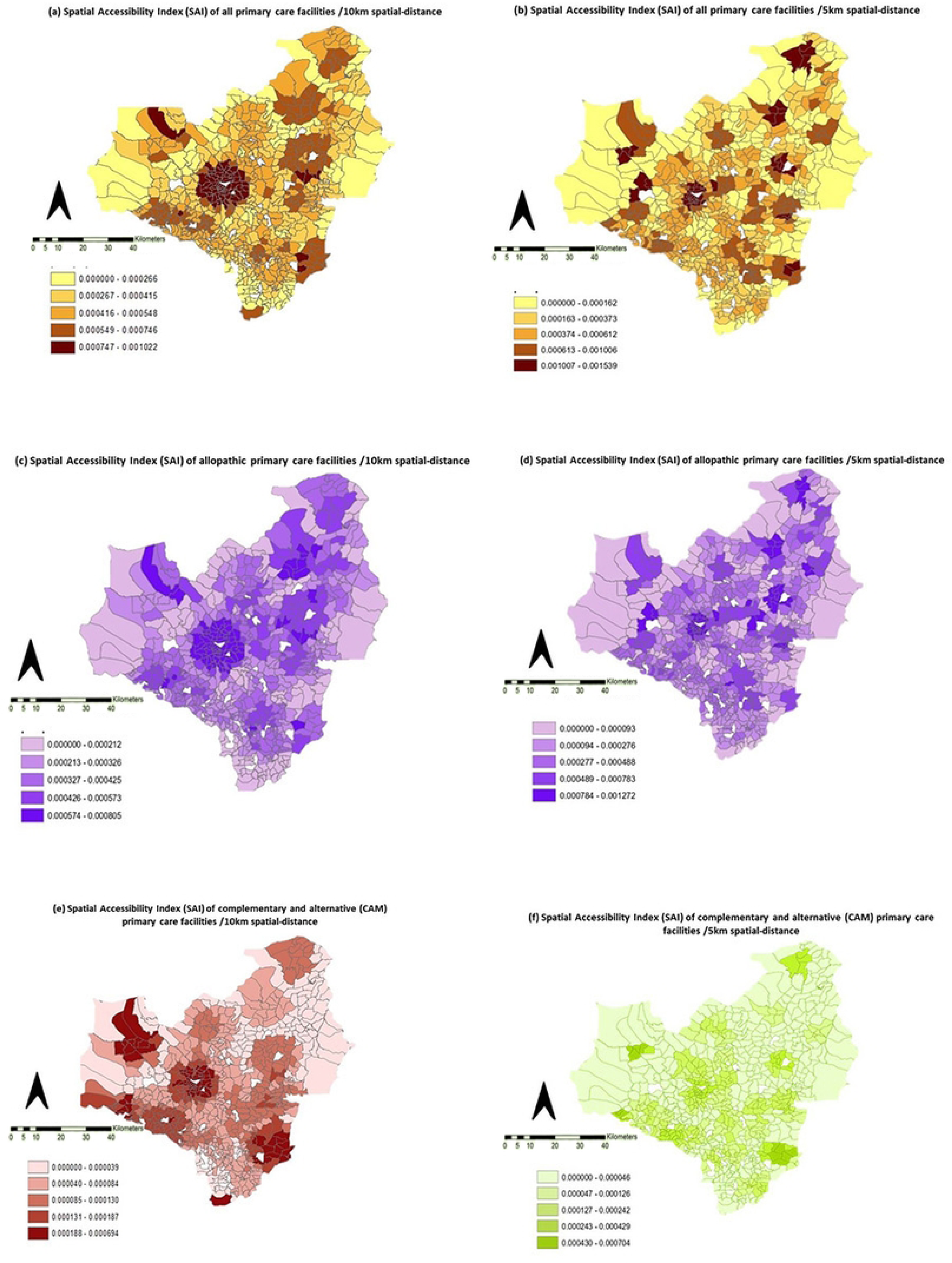
Spatial of distribution of Spatial Accessibility Index (SAI) of different types of primary care facilities in Anuradhapura district of Sri Lanka. The national target for primary care coverage (NPCC) of 1 PCD (if both allopathic and CAM are included) to 5000 population ratio was achieved in 70% (n=464) and 92% (n=605) of GND within 5 km and 10 km Spatial-distances. The coverage of 1 allopathic PCD to 5000 population ratio was achieved in 64% (n=420) and 86 % (n=565) GND within 5 km and 10 km Spatial-distances. Individual state and private allopathic PCD contributed 25% (n=166) and 56% (n=368) of GND to achieve NPCC in 10 km spatial-distance (Fig 3).

**Fig 3.**
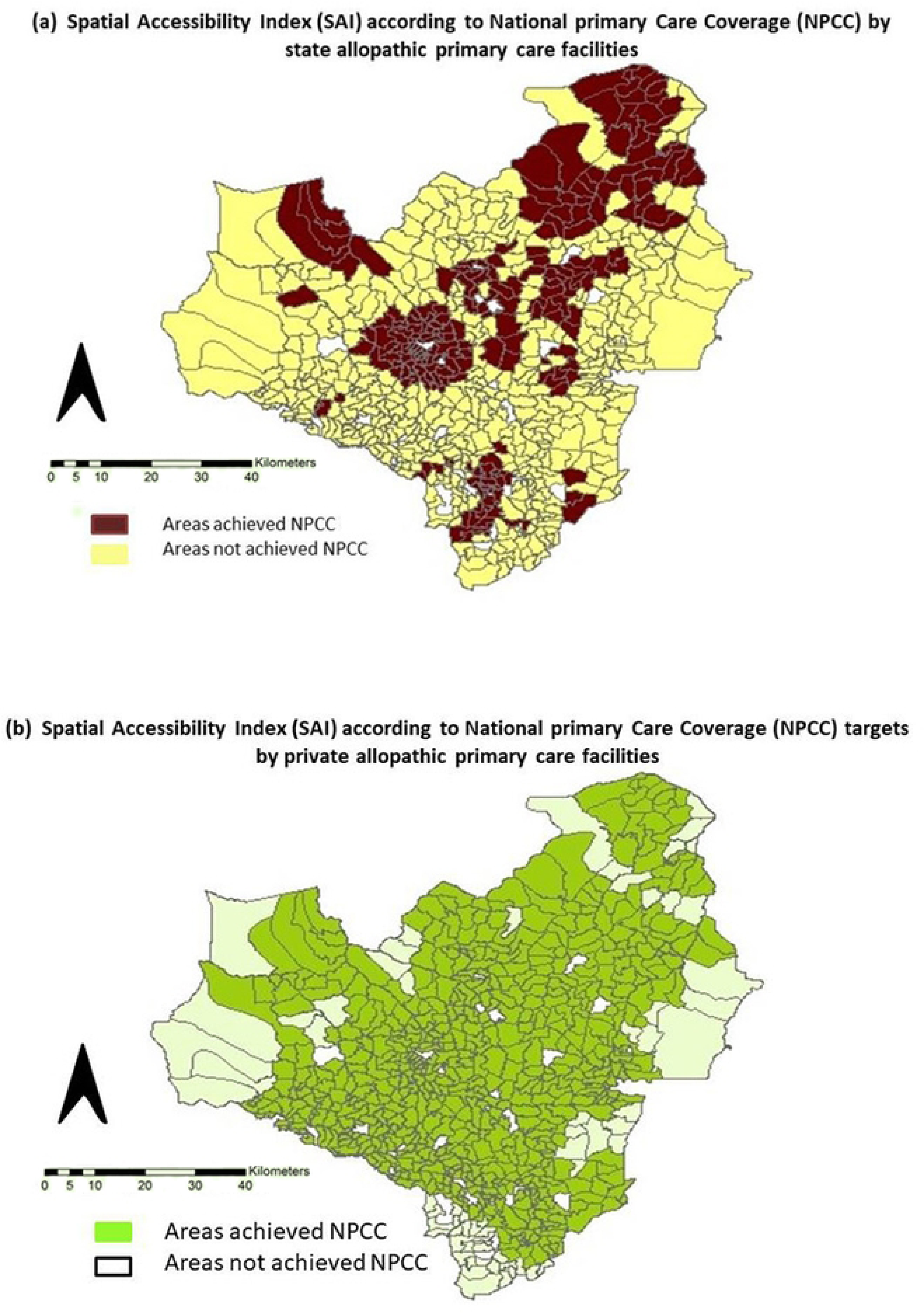
Spatial distribution of *Grama Niladhari* Divisions which achieved National Primary Care Coverage targets by state and private sector primary care doctors in Anuradhapura district of Sri Lanka. The difference between SAI values of the urban and rural areas was significant in 5 km and 10 km spatial-distance, p<0.005 (Independent sample t-test). The distance to the main road (r = -.218, p=0.0000, 5 km Spatial-distance) and the distance to main town, Anuradhapura (r= -.269, p=0.000, 5 km Spatial-distance) negatively correlated with SAI (S1 Table). SAI of divisional secretariat divisions within a 5 km Spatial-distance negatively correlated with poverty head count index (r= -.620, p=0.002). A positive correlation was observed between the mean SAI of divisional secretariat divisions (within 5 km Spatial-distance) and the population density, p< 0.001 (r = .754).

## Discussion

This study would be the first spatial methodological and GIS-based study in Sri Lanka to describe primary care spatial accessibility. It revealed how the private sector has expanded in both urban and rural populations as the main primary care provider. The spatial concentration of PCF was marked in urbanized areas compared to rural remotes. Despite the efforts for improving the human and physical resources related to PHC services in the country, it was alarming to find the gaps in rural service coverage and the relationship of accessibility with poverty indices and infrastructure facilities. The disparities in primary care access need to be recognized and evaluated by the policymakers for strategic planning to achieve 80% of UHC SCI by the year 2030. The spatial allocation of healthcare resources (23,24) with the use of SAI could be more helpful in low facility settings. One of the best evidence was provided from a Portuguese study and it demonstrated how the enhanced-2SFCA method was used to implement a new healthcare reform initiative for establishing emergency network points in a convenient distance to their residents (25).

Though the quality of service delivery at private hospitals was widely discussed in Sri Lankan health system-related studies (15,26) attribution of private allopathic PCD was overlooked due to lack of accurate databases (27). Although state healthcare staff was distributed proportionate to the patient capacity of the healthcare facilities, dismissal of spatial accessibility of the facilities has led to a disproportionate division of the healthcare workforce in the country. According to the study, only around one-third of the population in the district was located more than 5 km spatial-distance to reach their closest PCF. However, MoH reported that as high as 58% (28). The precise identification of spatial locations and comprehensive data collection greatly contributed to the accurate assessment of SAI through this study and it partially explains the observed difference between the surveys.

Based on the data, it is evident that the private sector was highly involved in the primary care workforce, accounting for 78% of it within the district. However the local reports highlighted an intriguing trend whereby households favoured utilizing state allopathic OPD (22%) as opposed to private allopathic PCF, 15% (29). When evaluating healthcare services utilization, it’s crucial to take into account the operating hours of PCF. More than just physical accessibility but affordability, accommodation, and accountability should be considered when comparing state and private PCF. The cost of private healthcare services in rural communities could potentially impose a financial burden on households (30), leading to limited access to healthcare.

The significant level of urban-rural disparity which observed in the study was reported in both LMIC (31,32) and HIC (33,34). The spatial concentration of PCD in urban areas was observed in this study due to two main factors: (1) the availability of tertiary and secondary care hospitals and (2) the higher availability of private allopathic practitioners. Several studies demonstrated how access to infrastructure and household income levels were associated with primary care spatial accessibility (24,35). Rural areas present significant challenges to retention of doctors due to various factors including inadequate physical infrastructure, limited educational opportunities, and lack of career development programs and earning opportunities. (36,37). The economic crisis in Sri Lanka could cause rural areas to lose doctors, further limiting healthcare access (38).

Within the district, the number of PCD to population ratio was reported as 3 for 20,000 population. Sri Lanka’s overall PCD to population ratio is unknown, but for the overall doctors from different levels of care (primary, secondary and tertiary), the ratio was 1:1000 (39). The total number of private allopathic PCD reported in the study was comparatively larger than the national reports of WHO (n=950) in 2021 (40). The unavailability of a proper GP registration system in the country is a main setback for accurate estimates of primary care workforces and predicting UHC SCI.

Intending to expand PHC services, the MoH has proposed a "shared care cluster system" as part of PHC reforms with an NPCC target of 1 PCD to a population of 5000 (14). Only 25% of the district population achieved NPCC via state-sector PCD, but the value reached 64% with the involvement of the private sector. A significant amount of remodeling of state PCF is needed to achieve the required primary care workforce in the district. The high level of private sector involvement raises the out-of-pocket expenses of the population. While regulating the quality of the private sector PCF, a coordinated collaboration between the state and private sector would be a timely strategy to achieve NPCC in the district and UHC SCI. A Proper assessment of equitable distribution of facilities should be a part of the healthcare facility planning process with feasible methods such as SAI spending minimal funds other than human efforts.

In low facility settings, transition from single PCD to primary care teams to primary care teams (41) and extension of telemedicine to remote areas (42,43) would improve primary care access for less expenditures.

### Limitations

MOH in the state sector was excluded from the study as most of his curative services were focused on specific areas in the preventive sector (e.g. maternal and child health) and not engaged in an open-extended commitment to primary care. Spatial maps which used in this study were last updated in year 2015 and the current boundaries and names of the GND might be different. The true SAI value may be different in some of the GND because of the edge effect (44) and geographical barriers (45) which were not considered in the analysis.

## Conclusion

The allopathic sector was the major primary care provider and, the private sector contributed to more than 75% of the primary care workforce in the Anuradhapura district. Ayurveda PCD provided one-fifth of the primary care workforce. Disparities in primary care access occurred due to socio-demographic factors such as the living location of the residents (rural or urban), infrastructure facilities, and the income level of the households. Nearly one-third of the population did not achieve NPCC requirements. Policy-level initiatives are needed to regulate the private sector, increase the retention of rural PCD and overcome urban-rural disparity in healthcare access. The spatial accessibility approach would be a feasible method to assess the equity of primary care access and resolve service gaps during healthcare planning.

## Data Availability

All relevant data are within the manuscript and its Supporting Information files.

## Acknowledgements

Technical support for ARC GIS analysis: PM Kankanamge, University of Sabaragamuwa, Sri Lanka and JMVA Jayawardhana Postgraduate Institute of Sciences, University of Peradeniya Sri Lanka.

## Author Contributions

Conceived and designed the study: PA, MW, SBA, SS, PHGJP. Funding acquisition for the study: PA. Supervision: PHGJP, SS, SBA. Performed the study: PA, MW, SBA, PHGJP. Analyzed the data: PA, SBA, SS, PHGJP. Contributed materials/analysis tools: PA, MW, SBA, PHGJP. Wrote the paper: PA, MW, SBA, SS, PHGJP.

## Supporting information

S1 Table Spatial Accessibility Index (SAI), proximity to closest primary care facilities and poverty data of Anuradhapura District of Sri Lanka

